# Diagnostic accuracy of three computer-aided detection systems for detecting pulmonary tuberculosis on chest radiography when used for screening: analysis of an international, multicenter migrants screening study

**DOI:** 10.1101/2022.03.30.22273191

**Authors:** Sifrash Meseret Gelaw, Sandra V. Kik, Morten Ruhwald, Stefano Ongarello, Tesfa Semagne Egzertegegne, Olga Gorbacheva, Christopher Gilpin, Nina Marano, Scott Lee, Christina R. Phares, Victoria Medina, Bhaskar Amatya, Claudia M. Denkinger

**Affiliations:** International Organization for Migration (IOM), Manila, Philippines; FIND, Geneva, Switzerland; International Organization for Migration (IOM), Geneva, Switzerland; United States Centers for Disease Control and Prevention (CDC), Atlanta, Georgia, United States; Heidelberg University Hospital, Center of Infectious Diseases, Heidelberg, Germany

**Keywords:** tuberculosis, screening, artificial intelligence, computer aided detection, diagnostic accuracy, chest-X-ray

## Abstract

The aim of this study was to independently evaluate the diagnostic accuracy of three artificial intelligence (AI)-based computer aided detection (CAD) systems for detecting pulmonary tuberculosis (TB) on global migrants screening chest x-ray (CXR) cases.

Retrospective clinical data and CXR images were collected from the International Organization for Migration (IOM) pre-migration health assessment TB screening global database for US-bound migrants. A total of 2,812 participants were included in the dataset, of which 1,769 (62.9%) had accompanying microbiological test results. All CXRs were interpreted by three CAD systems (CAD4TB v6, Lunit INSIGHT v4.9.0, and qXR v2) offline and re-interpreted by two expert radiologists in a blinded fashion. The performance was evaluated using receiver operating characteristics curve (ROC), estimates of sensitivity and specificity at different CAD thresholds against both microbiological and radiological reference standards (MRS and RadRS, respectively).

The area under the curve against MRS was highest for Lunit (0.85; 95% CI 0.83−0.87), followed by qXR (0.75; 95% CI 0.72−0.77) and then CAD4TB (0.71; 95% CI 0.68−0.73). At a set specificity of 70%, Lunit had the highest sensitivity (54.5%; 95% CI 51.7–57.3); at a set sensitivity of 90%, specificity was also highest for Lunit (81.4%; 95% CI 77.9–84.6). The CAD systems performed comparable to sensitivity (98.3%), and except CAD4TB, to specificity (13.7 %) of expert radiologist. Similar trends were observed when using RadRS.

In conclusion, the study demonstrated that the three CAD systems had broadly similar diagnostic accuracy with regard to TB screening, and comparable accuracy to expert radiologist. Compared with different reference standards, Lunit performed better than both qXR and CAD4TB against MRS, and better than qXR against RadRS. Overall, these findings suggest that CAD systems could be a useful tool for TB screening programs in remote, high TB prevalent places where access to expert radiologists may be limited.

## Introduction

Plain chest radiography remains a crucial tool for early detection of pulmonary tuberculosis (TB) and the monitoring of responses to TB treatment [1]. Chest X-rays (CXRs) have a high sensitivity in detecting pulmonary TB abnormalities, even in asymptomatic TB patients, especially when interpreted by experienced radiologists. Despite this, out of the estimated 10 million global TB cases in 2019, only 7.1 million were detected and reported [2]. As a result, although the global TB incidence rate and annual number of TB deaths has been steadily declining, it is not yet in line with the targets set out in the World Health Organization’s (WHO) End TB Strategy [2].

While advances in digital radiography technology have increased the quality of CXR images [3], limited access to these facilities and experienced radiologists remains a long-standing challenge, particularly in low-resource settings with a high TB burden [4]. However, recent advances in artificial intelligence (AI)-based computer-aided detection (CAD) systems have shown promising results in the automated interpretation of CXRs and detection of TB [5–7]. With acceptable accuracy, these CAD systems may help improve access to CXR reading for TB screening and contribute towards achieving WHO’s End TB strategy [2, 8]. However, there are limited number of studies in this area, most of which have methodological limitations, studied only one CAD software, with few screening data, and/or industry-funded, [7–10]. Moreover, most studies used non-expert CXR interpreters and assessed an online CAD processing system or shared images with the CAD vendors and compared the performance against a suboptimal refence standard of a single sputum specimen tested with Xpert MTB/RIF which further highlights the need for independent and rigorous studies [8–12]. More recent investigations have focused on offline and multiple AI systems [13–15], but they remain few in number.

A global consultation, convened by WHO in 2016, concluded that additional evidence on the performance and use of available CAD systems for TB screening were required [16]. To address this need, the International Organization for Migration (IOM) and FIND entered into a research collaboration to conduct two parallel studies at their respective organizations, both evaluating the accuracy of TB CAD technologies. The studies were conducted independently of the developers, using similar study designs and analysis plans, but involving separate archives. The results of both studies have contributed to the updated WHO consolidated guideline TB screening [17].

Here we present the results of the IOM study, evaluating the diagnostic accuracy of three commercially available CAD systems for detecting TB in offline setting using an independent global archive of CXR images collected from multiple TB screening of migrants in different countries. against a microbiological (MRS) and radiological reference standard (RadRS).

## Methods

### Study design

The manufacturer-independent archive of CXR images, set up at the IOM Global Teleradiology and Quality Control Center in Manila, consisted of retrospective clinical data and DICOM CXR images from multiple pre-migration health assessment TB screening of migrants bound for the US. These screenings were conducted across 31 IOM migrant health assessment Centers (MHACs) in 18 different countries between October 2014 and December 2017 (S1 and S2 Tables). For this study, all CXRs were analyzed by two experienced radiologists, as well as the three CAD systems.

### Study participants and screening assessments

The IOM Migration Health Division conducts pre-migration TB screening of refugees and immigrants bound to different resettlement countries through its various MHACs located in different countries worldwide. IOM uses a Global web-based application, Migrant Management Operation System Application (MiMOSA) to record migrants’ clinical information during the screening, and Local and Global Picture archive and communication systems (PACS) to archive the CXR images.

The TB screening of US-bound migrants is conducted in accordance with the US Centers for Disease Control and Prevention (CDC) Technical Instructions [18], which includes a clinical history, physical examination, and CXR examination (interpreted by qualified radiologist) using the standardized CXR reporting template provided by the US Department of State, DS-3030. Additionally, if the CXR reading is suggestive of TB or there is a clinical suspicion of TB, three consecutive sputum smear tests, plus solid and liquid culture tests, are completed. Molecular diagnostic tests such as Xpert are also performed if fast results are required or if there is a suspicion of drug-resistance. Participants eligible for inclusion in this study were 15 years or older, with a TB screening CXR for which the initial CXR interpretation and reference standards were available. No images included in this study had ever been shared with any of the CAD system manufacturers.

### Sample size and sampling

The sample size was calculated to demonstrate minimum CAD system accuracy targets of 90% sensitivity and 70% specificity, based on the WHO target product profile (TPP) for a TB triage test [19]. The minimum sample size required to detect these sensitivity and specificity targets with 90% power and a 95% confidence interval (CI) of 10% or less was 536 TB and 789 non-TB cases. These numbers were increased by 10% to account for missing information, resulting in a final target population of 590 TB and 868 non-TB cases.

Two samples were drawn from the screening archive (Sample 1 and Sample 2), one for each reference standard. Records were extracted from the MHACs with the highest caseloads first, until the sample size targets had been met.

### Data preparations

Biographic information, clinical and laboratory results, and original radiology readings were extracted from the IOM electronic Migrant Management Application, MiMOSA global database, and anonymized before being entered in the study dataset. DICOM CXR images of all participants were collected from each MHACs Local or Global PACS systems, as required, and also anonymized before further use in the study. Clinical and DICOM data were merged into one dataset using a unique participant identifier.

### Test methods (Index tests and Reference standards)

#### Index tests (CAD systems)

At the time of study initiation, the latest versions of commercially available, three “Conformité Européenne” (CE)-marked CAD systems that complied with European Union health, safety, performance, and environmental requirements were installed offline on an IOM-secured server: (1) CAD4TB version 6 (Delft Imaging, Netherlands; henceforth called CAD4TB); (2) Lunit INSIGHT CXR TB algorithm version 4.9.0 (Lunit INSIGHT, South Korea; henceforth called Lunit); and (3) qXR version 2 (Qure.ai, India; henceforth called qXR).

All three CAD systems read posterior-anterior (PA) CXR DICOM images and provide an abnormality score ranging from 0−100 (CAD4TB) or 0−1 (Lunit and qXR). A secondary image with a heatmap (CAD4TB and Lunit) or bounding boxes (qXR) is also produced that indicates the location of the identified abnormal findings (S1 Fig). Lunit and qXR have manufacturer-recommended thresholds for TB, while CAD4TB users are required to determine the threshold via a verification process (using data from the user site). Three threshold scores were provided by Lunit, either favoring high sensitivity (score=0.15), high specificity (score=0.45) or a middle threshold (score=0.3). Two thresholds were provided by qXR: a “routine TB screening threshold score” of 0.55 and a “high-risk TB threshold score” of 0.75.

For CAD4TB and qXR, a verification or test run was conducted on sets of CXRs from 13 types of different X-ray machine models used by IOM, which did not form part of this study, as required by the manufacturers at that time. Out of 13 tested, 10 X-ray machine models passed the verification for CAD4TB, and CXRs from those machines were included in the study (Agfa CR10-X, Agfa CR15-X, Agfa CR30-X, CareStream CR975, CareStream DRX-1, CareStream VitaCR, DRGEM, FUJIFILM, Kodak Point Of Care 260, SHIMADZU and SIEMENS). CAD CXR interpretation of DICOM images from study participants was carried out by IOM as per the manufacturer’s instructions, using offline server-installed CAD licenses. Only the PA CXR of the initial health assessment for each participant was used for the CAD interpretation, even if some participants had additional CXR views and follow-up CXRs.

#### Reference standards (microbiological [MRS] and radiological [RadRS])

For MRS analyses, a TB case was defined as a positive result on at least one out of three sputum cultures collected on consecutive days during the initial screening assessment. A non-TB case was defined as: (1) a negative result for all three sputum cultures, (2) the identification of non-tuberculous mycobacterium; and/or (3) at least one negative sputum culture result if the rest of the samples were contaminated. Only results from specimens taken within 14 days of the CXR were included. In the few cases where Xpert analyses were conducted, a positive Xpert result was interpreted as a positive MRS, even if the culture result was negative.

For RadRS analyses, all CXRs were analyzed by two certified IOM radiologists experienced in TB screening who had received specific training on TB screening CXR interpretation. The radiologists were blinded to the clinical and original CXR findings, as well as to the CAD results. Each specialist assessed half of the CXRs using the DS-3030 CXR reporting template containing a specified list of TB and non-TB findings (S3 Table). This re-assessment of the CXRs was conducted to reduce inter-reader variability and standardize the readings, as the original CXR interpretations were performed by several radiologists at different MHACs. If the image quality was not deemed to be acceptable, or if additional CXR views would have been required to complete the interpretation, the radiologists could exclude these CXRs from the analysis. When the new CXR readings showed major discrepancies with the original readings, CXR images were reviewed by a quality control radiologist, who provided a final reading after review of all interpretations from all sources. For RadRS, a TB case was defined as a CXR that was suggestive of active TB disease or old, healed TB (categories 2 and 3 of the CXR classification form; S3 Table). A non-TB case was defined as a normal CXR or one which showed other non-TB findings (categories 1, 4, 5, and 6; S3 Table).

### Data analysis

Clinical data, CXR readings, and CAD scores were collated into one dataset and any duplicates identified were excluded prior to analysis. Histograms of the CAD abnormality scores were plotted, receiver operating characteristic (ROC) curves calculated and the area under the curve (AUC) evaluated for each CAD system against both reference standards, using binomial distribution assumptions.

Estimates of sensitivity and specificity were also calculated at: (1) predefined points for sensitivity or specificity based on WHO triage TPP (90% sensitivity and 70% specificity); and (2) manufacturer-provided CAD score thresholds (only for Lunit and qXR) against MRS and RadRS. The sensitivity and specificity of radiologist assessments were also calculated against MRS. Finally, the sensitivity and specificity of each of the CAD systems against MRS were calculated at the threshold that produced the same specificity or sensitivity achieved by the radiologists.

Subgroup analyses for AUC, sensitivity and specificity were conducted for the following groups: age (15−35 years, 36−55 years, 56+ years), sex, geographical region, high-risk groups (e.g., a history of previous TB), migrant type (refugee vs immigrant), HIV status (if known), presence of TB symptoms, sputum smear status, presence of some image quality issues even if the images were deemed acceptable overall, and the presence of additional CXR views obtained during the screening and re- assessed by expert radiologists. Stata® version 16 was used for data management and analysis [20].

### Ethical considerations

The study was conducted after receiving all appropriate approvals, legal agreements, and authorizations, and in accordance with all applicable data privacy and security rules. The study received IOM legal council approval to use participant’s data and CDC approval was obtained for use of data from the pre-migration health assessment program. The study protocol received ethical approval from McGill University Health Centre Institutional Review Board (project number 2019-4649).

## Results

### Study selection

A total of 2,910 cases were sampled (Fig 1): 589 culture-positive and 865 culture-negative from Sample 1, and 590 CXR suggestive of TB and 866 CXR not suggestive of TB from Sample 2. After 78 duplicates were excluded and 20 cases were rejected by the expert radiologists based on the CXRs, 2,812 cases were included in the RadRS analysis. Of these, 1,769 were also included in the MRS analysis. The remaining 1,043 cases were not used for analysis against MRS because either they had > 14 days between the CXR exam and sputum collection or there were no available sputum culture or Xpert results. Additionally, 207 participants with invalid CAD4TB scores (empty or negative values) were excluded from analysis for CAD4TB; Lunit and qXR had valid scores for all observations.

**Fig 1.**
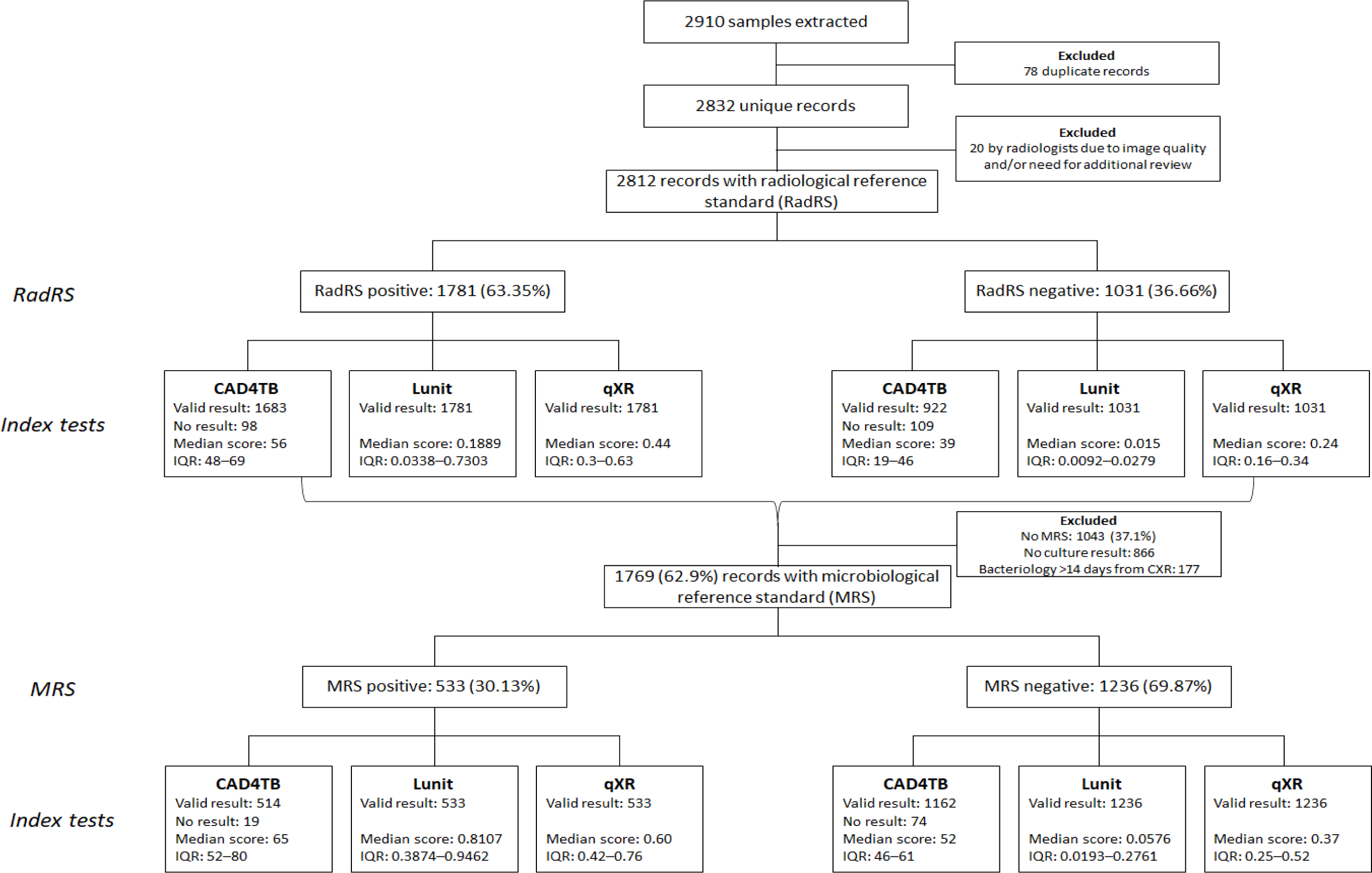
Flowchart of participants included in the analysis.

### Baseline demographic and clinical characteristics

Baseline demographic and clinical characteristics of the whole study population (RadRS) and the population included in the analysis against MRS are presented in S4 Table and Table 1, respectively. While CXR findings and microbiological test results among RadRS and MRS population are presented in S5 Table and Table 2, respectively.

**Table 1.** Baseline demographics and clinical characteristics (MRS analysis population).

| Variables | TB (%) | non-TB (%) | Total in MRS (%) |
| --- | --- | --- | --- |
| **Total** | 533 (30.1%) | 1236 (69.9%) | 1769 (100%) |
| Male | 357 (67.0%) | 714 (57.8%) | 1071 (60.5%) |
| Female | 176 (33.0%) | 522 (42.2%) | 698 (39.5%) |
| **Age group** |  |  |  |
| 15–35 year | 237 (44.5%) | 404 (32.7%) | 641 (36.2%) |
| 36–55 year | 166 (31.1%) | 395 (32.0%) | 561 (31.7%) |
| 56+ year | 130 (24.4%) | 437 (35.4%) | 567 (32.1%) |
| **Region** |  |  |  |
| Africa | 125 (23.5%) | 305 (24.7%) | 430 (24.3%) |
| Asia Pacific | 391 (73.4%) | 792 (64.1%) | 1183 (66.9%) |
| Middle East | 2 (0.4%) | 15 (1.2%) | 17 (1%) |
| Eastern Europe | 14 (2.6%) | 124 (10.0%) | 138 (7.8%) |
| not indicated | 1 (0.2%) | 0 (0.0%) | 1 (0.1%) |
| **TB symptoms** |  |  |  |
| Cough (any duration) | 15 (2.8%) | 5 (0.4%) | 20 (1.1%) |
| Fever | 5 (0.9%) | 1 (0.1%) | 6 (0.3%) |
| Weight loss | 11 (2.1%) | 3 (0.2%) | 14 (0.8%) |
| Night sweats | 5 (0.9%) | 0 (0.0%) | 5 (0.3%) |
| TB symptoms present, $\geq 1$ | 19 (3.6%) | 8 (0.6%) | 27 (1.5%) |
| **Risk groups** |  |  |  |
| History of TB | 99 (18.6%) | 152 (12.3%) | 251 (14.2%) |
| Migrant type (refugee) | 325 (61.0%) | 692 (56.0%) | 1017 (57.5%) |
| HIV positive | 12 (2.3%) | 6 (0.5%) | 18 (1%) |
| **Image quality issue** |  |  |  |
| Yes | 103 (19.3%) | 208 (16.8%) | 311 (17.6%) |
| No | 430 (80.7%) | 1028 (83.2%) | 1458 (82.4%) |
| **Additional view present** |  |  |  |
| Yes | 79 (14.8%) | 319 (25.8%) | 398 (22.5%) |
| No | 454 (85.2%) | 917 (74.2%) | 1371 (77.5%) |
| **Smear result** |  |  |  |
| Positive | 159 (30.0%) | 16 (1.3%) | 175 (9.9%) |
| Negative | 371 (70.0%) | 1220 (98.7%) | 1591 (89.9%) |

**Table 2:** Chest X-ray findings and microbiological test results (MRS analysis population).

| **Variables** | **TB (%)** | **non-TB (%)** | **Total in MRS (%)** |
| --- | --- | --- | --- |
| **Total** | 533 (30.1%) | 1236 (69.9%) | 1769 (100%) |
| **CXR finding result** |  |  |  |
| 1=Normal | 6 (1.1%) | 114 (9.2%) | 120 (6.8%) |
| 2=Abnormal CXR, highly suggestive of active TB (follow-up required) | 495 (92.9%) | 686 (55.5%) | 1181 (66.8%) |
| 3=Abnormal CXR, may suggest old, healed TB but active TB can't be ruled out (follow-up required) | 24 (4.5%) | 359 (29.1%) | 383 (21.7%) |
| 4=Abnormal CXR, can remotely suggest old, healed TB but minimal risk of reactivation (NO follow-up required) | 3 (0.6%) | 53 (4.3%) | 56 (3.2%) |
| 5=Abnormal CXR, not suggestive of TB (follow-up required) | 5 (0.9%) | 24 (1.9%) | 29 (1.6%) |
| 6=Other abnormalities (NO follow-up required) | 0 (0.0%) | 0 (0.0%) | 0 (0%) |
| **Total** | 533 (100%) | 1236(100%) | 1769 (100%) |
| **Bacteriology result (for those done =<14 days of CXR to culture date)** |  |  |  |
| Culture +, Xpert MTB/RIF not done | 450 (84.4%) | 0 (0.0%) | 450 (25.4%) |
| Culture +, Xpert MTB/RIF + | 81 (15.2%) | 0 (0.0%) | 81 (4.6%) |
| Culture +, Xpert MTB/RIF - | 1 (0.2%) | 0 (0.0%) | 1 (0.1%) |
| Culture -, Xpert MTB/RIF + | 1 (0.2%) | 0 (0.0%) | 1 (0.1%) |
| Culture -, Xpert MTB/RIF not done | 0 (0.0%) | 1228 (99.4%) | 1228 (69.4%) |
| Culture -, Xpert MTB/RIF - | 0 (0.0%) | 8 (0.7%) | 8 (0.5%) |
| **Total** | **533 (100%)** | **1236 (100%)** | **1769 (100%)** |

In the MRS analysis population, more than half (60.5%) were male, most were young (36.2% were 15−35 years of age), and 30.1% were MRS positive. MRS-positive TB cases were reported more often among males (67.0%), at a younger age (44.5% were 15–35 years of age), and in those with TB symptoms compared to non-TB cases (3.6% vs 0.6%). Sputum smears were positive in 29.8% of MRS- positive cases, and abnormal CXR findings in 99%, 97.5% of which were CXR suggestive of TB (Table 1). However, 91% of the non-TB cases also had abnormal CXR findings, 84.6% of which were CXR suggestive of TB. Only 4.7% of MRS TB cases had Xpert results in addition to cultures, and only two (0.1%) had discrepancies, one being culture-negative and Xpert-positive, and the other being culture- positive and Xpert-negative (Table 2). In the RadRS analysis population, 63.3% had CXR suggestive of TB were RadRS positive cases. Of those, 32.3% were culture positive (S5 Table).

### Histogram distribution of index tests

Abnormality scores of all three CAD systems showed some bimodal distribution when plotted in a two-way histogram against MRS, with a wide range of overlap between TB and non-TB cases (S2a Fig). Of all CAD systems, Lunit provided the least overlap. Similar distributions were observed in the analysis against RadRS (S2b Fig).

### Overall diagnostic accuracy of index tests

The AUCs of the ROC curves, plotting the estimated sensitivity and specificity at each possible abnormality score, (using MRS) were highest with the Lunit system (0.85; 95% CI 0.83−0.87), followed by qXR (0.75; 95% CI 0.72−0.77) and then CAD4TB (0.71; 95% CI 0.68−0.73) (Fig 2a). Expert radiologist analysis of the CXRs for TB showed a sensitivity of 98.3% (95% CI 96.8−99.2%) and specificity of 13.7% (95% CI 11.8−15.7) against the same MRS (Fig 2a).

**Fig 2:**
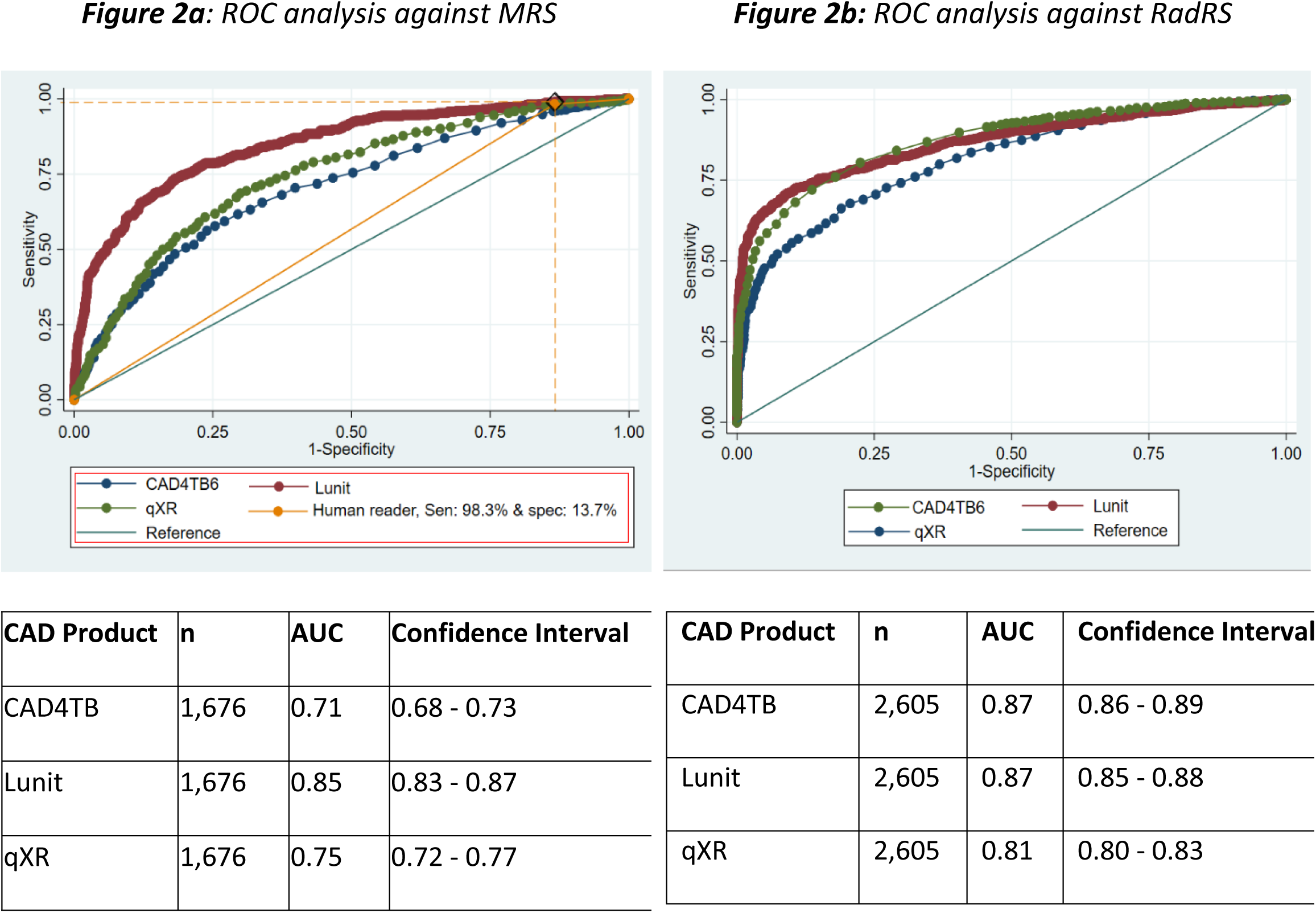
Receiver operating characteristic curves of three CAD systems against microbiological and radiological reference standards. Cases with no valid CAD4TB scores were excluded from the ROC analysis (n=93 from the ROC against MRS; n=207 against RadRS).

The ROC curves against RadRS showed greater AUCs for all CAD systems compared with the MRS analysis, being highest with CAD4TB (0.87; 95% CI 0.86−0.89) and Lunit (0.87; 95% CI 0.85−0.88), followed by qXR (0.81; 95% CI 0.80−0.83; Fig 2b).

### Accuracy estimates of index tests

The sensitivity and specificity of the three CAD products computed at a range of thresholds against MRS showed the highest accuracy (a combination of sensitivity and specificity) for Lunit in all target categories, followed by qXR and CAD4TB. At a set sensitivity of 90%, specificity values were 54.5% (95% CI 51.7−57.3%) for Lunit, 32.4% (95% CI 29.8−35.1%) for qXR, and 23.0% (95% CI 20.6−25.5%) for CAD4TB. At a set specificity of 70%, sensitivity values were 81.4% (95% CI 77.9−84.6%) for Lunit, 67.9% (95% CI 63.8−71.9%) for qXR, and 61.7% (95% CI 57.3−65.9%) for CAD4TB.

At the sensitivity value achieved by expert radiologists (98.3% against MRS), the specificities of Lunit and qXR were 15.8% (95% CI 13.8−17.9%) and 12.0% (95% CI 10.2−13.9%), respectively, compared to 13.7% (95% CI 11.8−15.7) for the expert radiologist. CAD4TB had a lower specificity of 6.5% (95% CI 5.2−8.1%). At the specificity value achieved by expert radiologists (13.7% against MRS), the sensitivity was highest for Lunit (99.1%; 95% CI 97.8−99.7%), followed by qXR (97.7%; 95% CI 96.1−98.8%), then CAD4TB (95.9%; 95% CI 93.8−97.5%; Table 3), compared to 98.3% (95% CI 96.8−99.2%) for the expert radiologist.

**Table 3:**
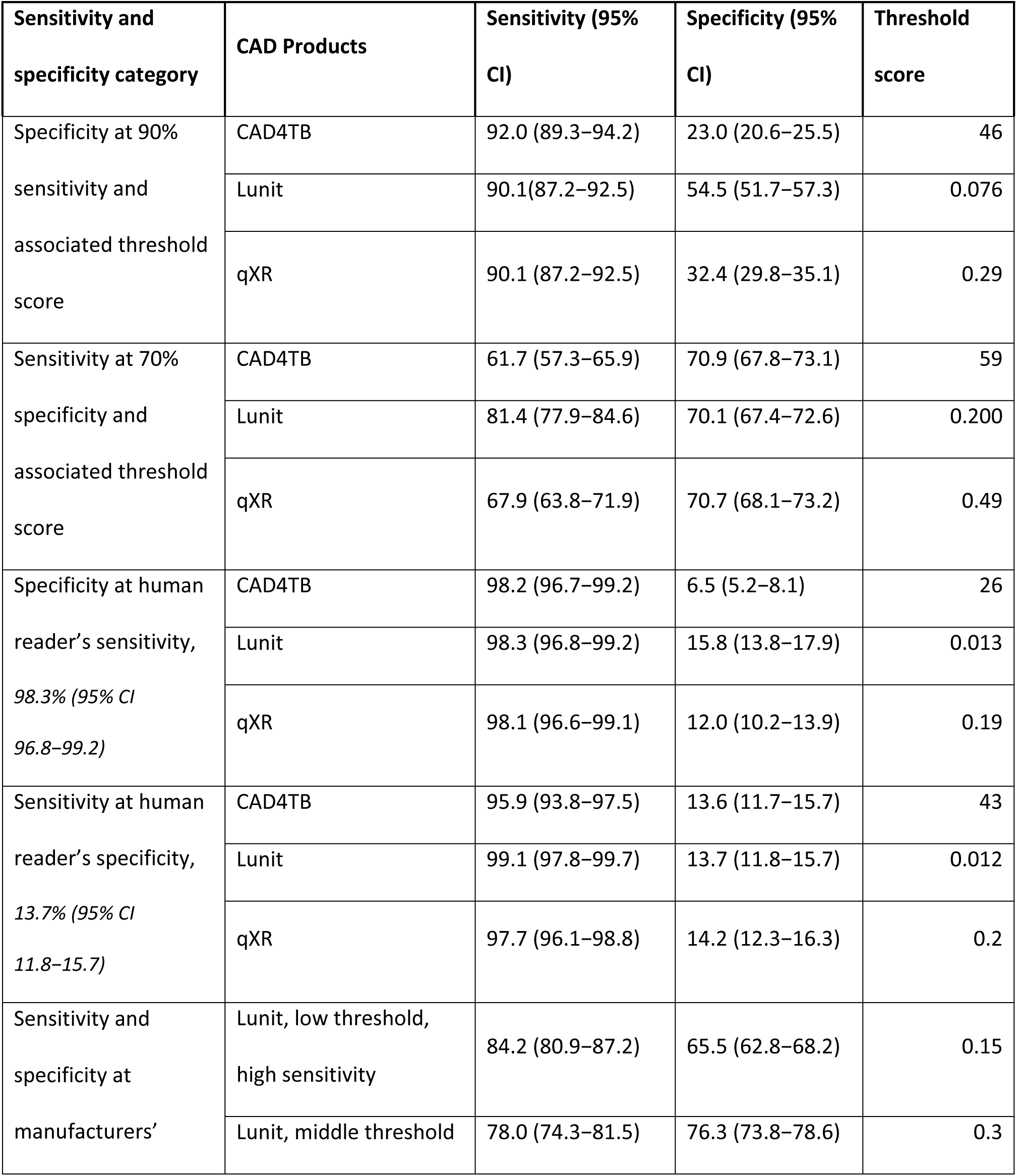
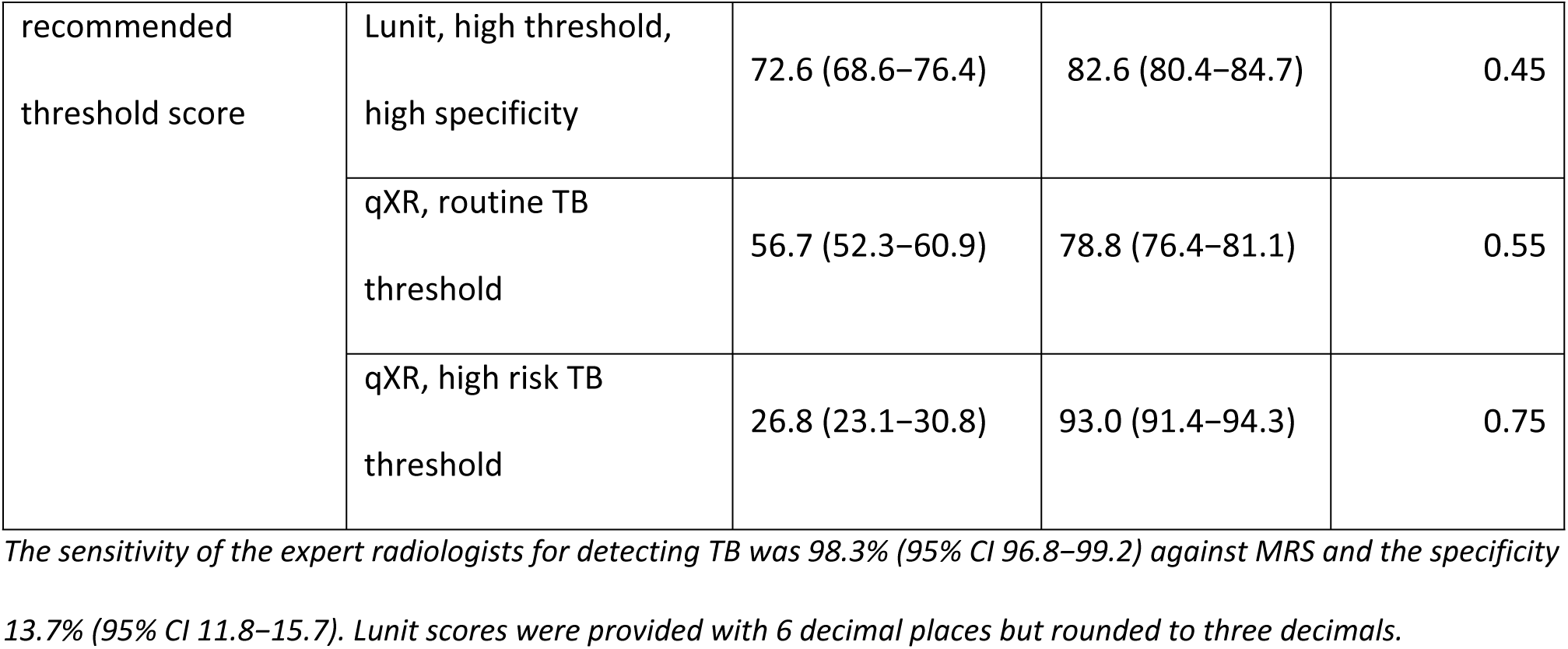
Accuracy estimates of index test at different sensitivity and specificity points (MRS analysis population).

At manufacturer-recommended thresholds, the three Lunit thresholds (0.15, 0.3 and 0.45) resulted in sensitivities of 84.2% (95% CI 80.9−87.2), 78.0% (95% CI 74.3−81.5) and 72.6% (95% CI 68.6−76.4), with specificities of 65.5% (95% CI 62.8−68.2), 76.3% (95% CI 73.8−78.6), and 82.6% (95% CI 80.4−84.7) respectively. However, the targets of the WHO TPP for a triage test were not met at any of the thresholds. For qXR, both threshold scores (0.55 and 0.75) resulted in lower sensitivities than Lunit: 56.7% (95% CI 52.3−60.9) and 26.8% (95% CI 23.1−30.8), with specificities of 78.8% (95% CI 76.4−81.1) and 93.0% (95% CI 91.4−94.3 respectively; Table 3).

Sensitivity and specificity estimates against RadRS are presented in S6 Table. At 90% sensitivity, specificity was highest for CAD4TB, followed by Lunit and then qXR. At 70% specificity, CAD4TB and Lunit had a higher sensitivity than qXR (S6 Table). At the manufacturer-recommended thresholds, both Lunit and qXR showed similar trends to the analysis against MRS, but typically with lower sensitivity and higher specificity values.

Image processing errors were noticed for 178 CXR images after processing by Lunit, in which the images were inverted from the original negative image (bones white) to positive (bones black). The sensitivity and specificity values, as well as the AUC of Lunit with and without those cases included, were unaffected by these processing errors.

### Diagnostic accuracy of index tests in different population subgroups

The diagnostic accuracy, expressed in terms of the AUC of the ROC curve for all three CAD systems, was lower in cases with a history of pulmonary TB compared to those without a history of TB, and lower with CAD4TB in smear-negative cases (0.66; 95% CI 0.63−0.69), compared to smear-positive cases (0.82; 95% CI 0.70−0.94) and in those with additional views (0.58; 95% CI 0.50−0.65), compared to those without additional views (0.72; 95% CI 0.69−0.75) (S3 Fig and S7 Table). For other subgroups such as female sex, HIV infected, absence of TB symptoms, and immigrants (and for Lunit the older age group), CAD systems appeared to show lower AUC estimates compared with their opposing subgroups, though the CIs overlapped. Other subgroups, such as image quality and region, did not show any additional trends (S3 Fig and S7 Table).

## Discussion

This study is one of the first comprehensive studies evaluating CAD systems independent of the CAD developers in a population screened for TB using both culture results and expert radiologist assessments as reference standards. The findings from the study demonstrated that the three CAD systems (Lunit, CAD4TB, qXR) have comparable diagnostic accuracy in detecting TB on CXR when used for TB screening and may perform comparably to that of expert radiologists, with Lunit performing better than both qXR and CAD4TB against MRS and CAD4TB and Lunit performing better than qXR against RadRS. However, none of the CAD systems reached the minimum performance requirements of the WHO triage TPP (90% sensitivity and 70% specificity) [19], in contrast to previously published findings by Khan et al. [13].

The finding that (i) Lunit performed best against MRS and (ii) CAD4TB performed best against RadRS, shows that the CADs performance can vary by the reference standard used, and may indicate that Lunit is better at detecting CXR findings suggestive of active TB disease, which tend to be culture- positive, while CAD4TB may better detect CXR findings suggestive of old, healed TB that can be identified by radiologists, but tend to be culture-negative. This finding could also reflect the methodology used in the deep machine learning of the CAD product algorithms, e.g., mainly training the software against RadRS versus MRS. The better performance of CAD4TB against RadRS than MRS is also supported by the results of a previous study by Fehr et al. [11].

The low specificity of the CADs at a set sensitivity of 90% against MRS is similar to the expert radiologist and could also be explained by the intrinsic nature of sputum culture, CXR, and CXR signs of TB. Culture analysis detects TB in cases with detectable bacteria in the sputum. As such, it measures the sensitivity of detecting active TB disease, whereas, old, healed pulmonary lesions detected by CXR can be culture-negative and are considered false-positive in the analysis against MRS. Additionally, CXR signs of TB are not specific to TB only, thereby reducing the estimated specificity. Therefore, CXR is recommended for screening but not as a confirmatory diagnostic tool (i.e., a positive CXR TB screening result should be used as criteria for further confirmatory testing, such as sputum cultures, and not for a treatment decision). Nevertheless, for a screening tool the benefit of high sensitivity may outweigh the limitations of a lower specificity. Both Lunit and qXR had relatively lower sensitivity and specificity at all manufacture provided thresholds, though Lunit performed with relatively higher sensitivity while qXR achieved higher specificity. As such, the sensitivity and specificity thresholds of the CADs that correspond with expert radiologist assessments (98.3% and 13.7%, respectively), may be potential candidates for the selection of optimal thresholds for operational use.

Subgroup analyses showed that the performance of CADs can vary among some population demographic and clinical characteristics. All CAD systems performed worse in participants with a history of TB, something which has also been observed in previous studies [13–15]. This is to be expected, as healed TB can leave residual CXR changes, which usually are classified as TB findings on CXR but can lead to negative microbiological test results. CAD4TB performed lower in participants with smear-negative results, in line with the findings of Khan et al. [13]; CAD4TB, moreover, performed worse in cases where additional CXR views were requested by the expert radiologist. Again, these results are not surprising, as smear-positive cases may have obvious CXR abnormalities that can be easily detected, and the absence of a request for additional views may indicate that the initial CXR was of good quality and/or there were no suspicious CXR findings. However, this conclusion was significant only for CAD4TB, while a similar although not statistically significant trend was observed for Lunit and qXR.

Additional trends were observed in the other subgroup analyses. While overlapping CI values indicate that these findings should be interpreted with caution, it appeared that the CAD systems performed less well in females, participants with no TB symptoms, HIV-positive participants, those with an ‘immigrant’ status compared with those classified with a ‘refugee’ status, and in older participants (for Lunit only). Other studies have also reported that CAD performance can be significantly impacted by sex and age [13]. The differences in CAD performance among different subgroups indicates that population characteristics should be taken into consideration before implementation.

There are some limitations to this study that should also be considered. Firstly, participants received sputum smear and culture tests during the initial TB screening only when the initial CXR reading was suggestive of TB or there was a clinical suspicion of TB. Therefore, sputum culture testing was not performed for most participants with normal CXRs or CXR findings suggestive of non-TB and were not included in the MRS analysis. This likely resulted in lower specificity readings for the CADs and expert radiologists, as would be expected from an unselected population. Moreover, as TB cases were overrepresented for both the MRS and RadRS analyses due to the sampling strategy employed in this study, the dataset may not be representative of all people presenting for TB screening but is instead a subset of those who had a higher suspicion of TB and therefore underwent sputum examination.

This might have affected the overall accuracy estimates, albeit to a similar extent for all three CAD systems and the expert radiologists. Additionally, 20 participants with no radiologist assessment were excluded from the analyses, as well as 207 images from the CAD4TB analysis due to invalid score results. The reason for the invalid scores with the CAD4TB system was unknown, but it could be because the software quality control rejected unacceptable or poor images without requiring further investigation. Although the number of excluded results is small compared with the size of the whole dataset, it is possible that the characteristics of the cases excluded may have been different from those that were included.

Although the study did not systematically evaluate quality control measures of the CADs, some issues were observed during the automated interpretation of the CXRs by the CAD systems. Some CXR projections other than the PA CXRs, such as lateral and lordotic CXR views (which are unsupported by the CADs) or CXRs with image quality issues, were not always flagged by the systems. The study also did not assess the operational performance of the CADs such as the processing time, technical issues, and troubleshooting responses, infrastructure needs, comparison of offline and online use of the CAD product, cost-effectiveness, or other related matters. In addition, since the study was conducted new versions of the CAD systems have been released and other CAD systems have entered the market [21], which may necessitate further evaluation.

Based on the findings of this study, combined with those of the parallel study conducted by FIND [22], CAD systems may be considered viable as a tool for automated CXR interpretation with regard to TB detection in screening programs, particularly in remote, and/or a high TB burden places where there are limited resources and access to expert radiologists. The use of CAD systems in these areas may even have a wider application and contribute to increase the global TB detection rate. Further to these, and other, findings, WHO has recently released consolidated guidelines on tuberculosis recommending that CAD may be used in place of human readers for interpreting digital CXR for TB screening in individuals aged 15 years and older [17]. Another role of CADs, even in places where expert radiologists are available, may be their use for internal quality control monitoring of CXRs complementary to radiologist assessments.

Nevertheless, further studies may be required to further investigate the accuracy of CADs in detecting non-TB-significant findings, such as lung cancer or bone lesions as well as the different specific CXR findings suggestive of TB, better address the performance of CADs in the different population subgroups, the way the CADs address image quality issues that might necessitate repeat CXRs or additional views by radiologists, how the CADs handle non-PA CXRs, and non-complied age requirements for specific systems. Likewise, for operational use of the CAD systems, additional questions should be addressed [23], including choice of the CAD system and version, compatibility with X-ray machine, accepted image format, need for validation, integration into existing workflow and patient registration systems, feasibility of online or offline use of the software, and technical requirements, as well as the selection of optimal thresholds for the intended use.

In conclusion, the results of this study demonstrated the comparability of the accuracy of three CAD systems for CXR interpretation with regard to TB screening, which may broadly perform similar to that of an expert radiologist. Additionally, the study has demonstrated that the performance of the CAD systems can vary by population demographic and clinical characteristics as well as the reference standard used. As such, these tools may provide viable options for use in TB screening programs to increase TB detection, especially in low resource areas where there may be no available expert radiologists. However, further studies are needed to better address CAD performance in specific population subgroups or different CXR TB findings, to assess other operational and technical factors necessary for proper operational implementation, and to evaluate novel CAD products coming to the market.

## Data Availability

The data used for this study was obtained from IOM?s pre-migration health assessment of migrants bound for the United States after getting approval from the IOM legal counsel and CDC for the purposes of this study only. The IOM data protection policy restricts sharing data with any third party.

## Acknowledgments

We would like to thank all the study participants who took part in the IOM health assessment screening and included in the study, the three CAD software companies, Delft Imaging, Qure.ai and Lunit INSIGHT, for installing the CAD products on the IOM server and providing the technical support and troubleshooting, CDC for approving the use of the data and CXR images from the IOM health assessment TB screening of migrants bound to the United States, and Dr. Mary Naughton and Dr. Drew Posey from CDC for assisting in the process, Dr. Cecily MILLER and Dr. Dennis FALZON from the WHO TB programme for their technical collaboration during the study and allowing us to present results of the studies to the WHO TB guideline development group meeting, FIND for getting funding for the project and technical coordination, IOM Legal counsel for approving to use the Data and CXR images of migrants, as well as Dr Fiaz Ahmad Khan from McGill university for his coordination of the Ethical approval. Additionally, we thank our IOM colleague Mr. Rommel Cordero for assisting the technical support and troubleshooting, our IOM Teleradiology Radiologists Dr. Ethel Enriquez-Alas, Dr. Sthennette Jerusalem for re-interpreting the CXRs included in the study, Dr. Lena Maria Ablis-Sun for assisting in quality control review of the discrepant cases and finalizing the CXR report, Ms. May Antonnette Lebanan for her technical guidance during the data analysis, and Dr. Paul Douglas, and Mrs. Jacqueline Weekers and Mr. Enrico Ponziani for the overall support and guidance.

## Disclaimer

The findings and conclusions of this article are those of the authors and do not necessarily represent the official position of the US Government or CDC. References in this manuscript to any specific commercial CAD products, process, service, manufacturer, or company do not constitute endorsement or recommendation by the US Government or CDC.

## Related manuscripts

This study done at IOM is one of the two studies on accuracy of CAD for screening use cases conducted at IOM and FIND under the same umbrella study protocol but using different data sources and methods. The study conducted at FIND is sent for publication separately.

## Authors’ contributions

SMG, SK, and CMD contributed to the conception of the study, writing the study protocol, SMG and SK contributed to the whole process of the study from sampling, data extraction, data analysis, data interpretation and updating the manuscript. SMG contributed leading the overall process of this study and drafting the manuscript. CMD, MR contributed to data interpretation. SO contributed to the study protocol, the statistical analysis plan and the data analysis. OG reviewed the Proposal and assisted in the legal and ethical approval. CG assisted in funding acquisition, TSE assisted in the data reporting and monitoring of the re-reading of the CXRs by expert radiologists. NM and CP reviewed the study protocol, assisted CDC approval to use the data, BA assisted in technical CAD set up, CXR image processing, and data collection, VM assisted in data analysis. SL and all co-authors reviewed and agreed on the final manuscript.

## Data availability

The data used for this study was obtained from IOM’s pre-migration health assessment of migrants bound for the United States after getting approval from the IOM legal counsel and CDC for the purposes of this study only. The IOM data protection policy restricts sharing data with any third party.

## Supporting information

**S1 Fig:**
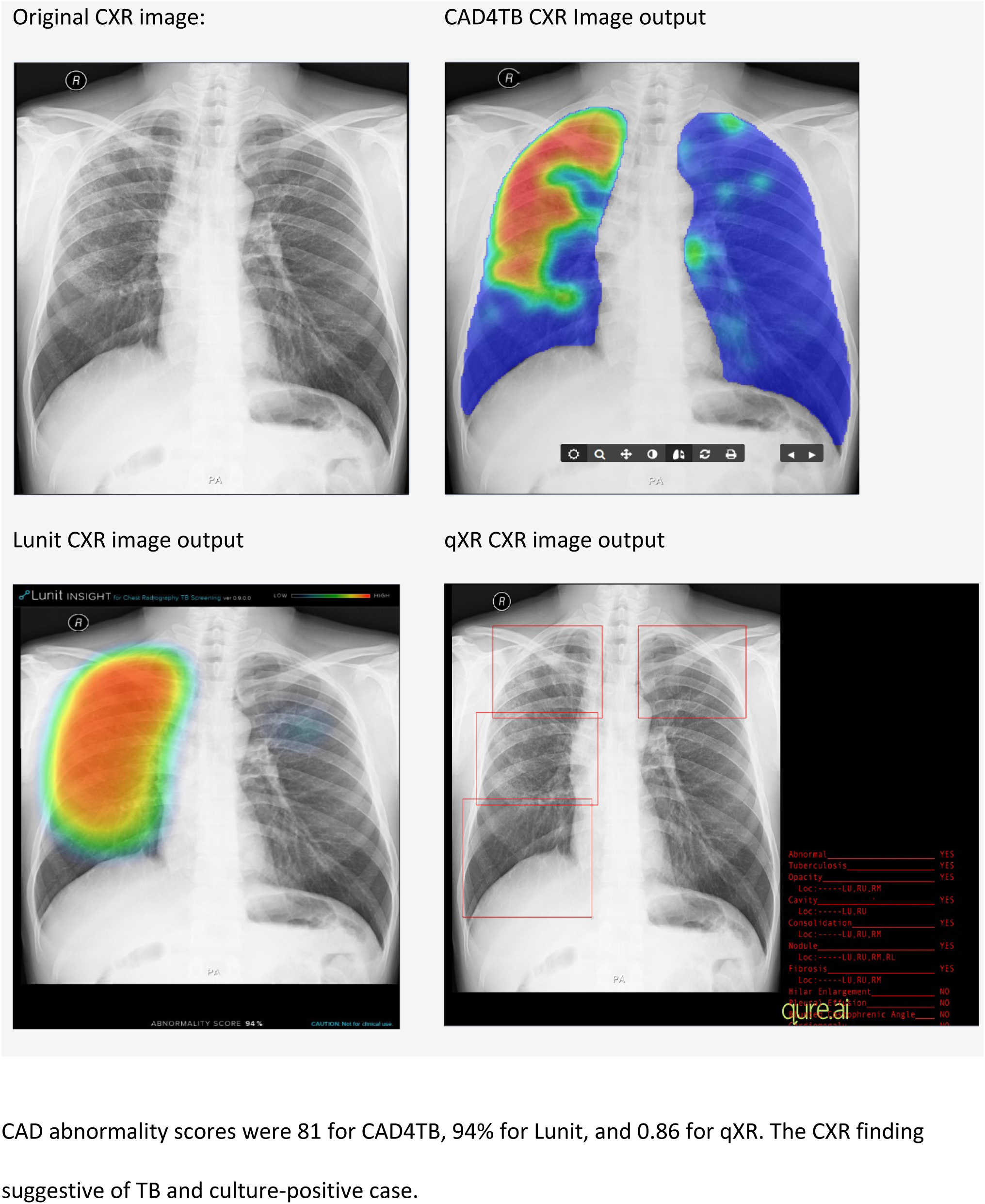
Sample chest x-ray image before and after the image processing by each CAD system, with image output heat maps/ boxes. CAD abnormality scores were 81 for CAD4TB, 94% for Lunit, and 0.86 for qXR. The CXR finding suggestive of TB and culture-positive case.

**S2 Fig:**
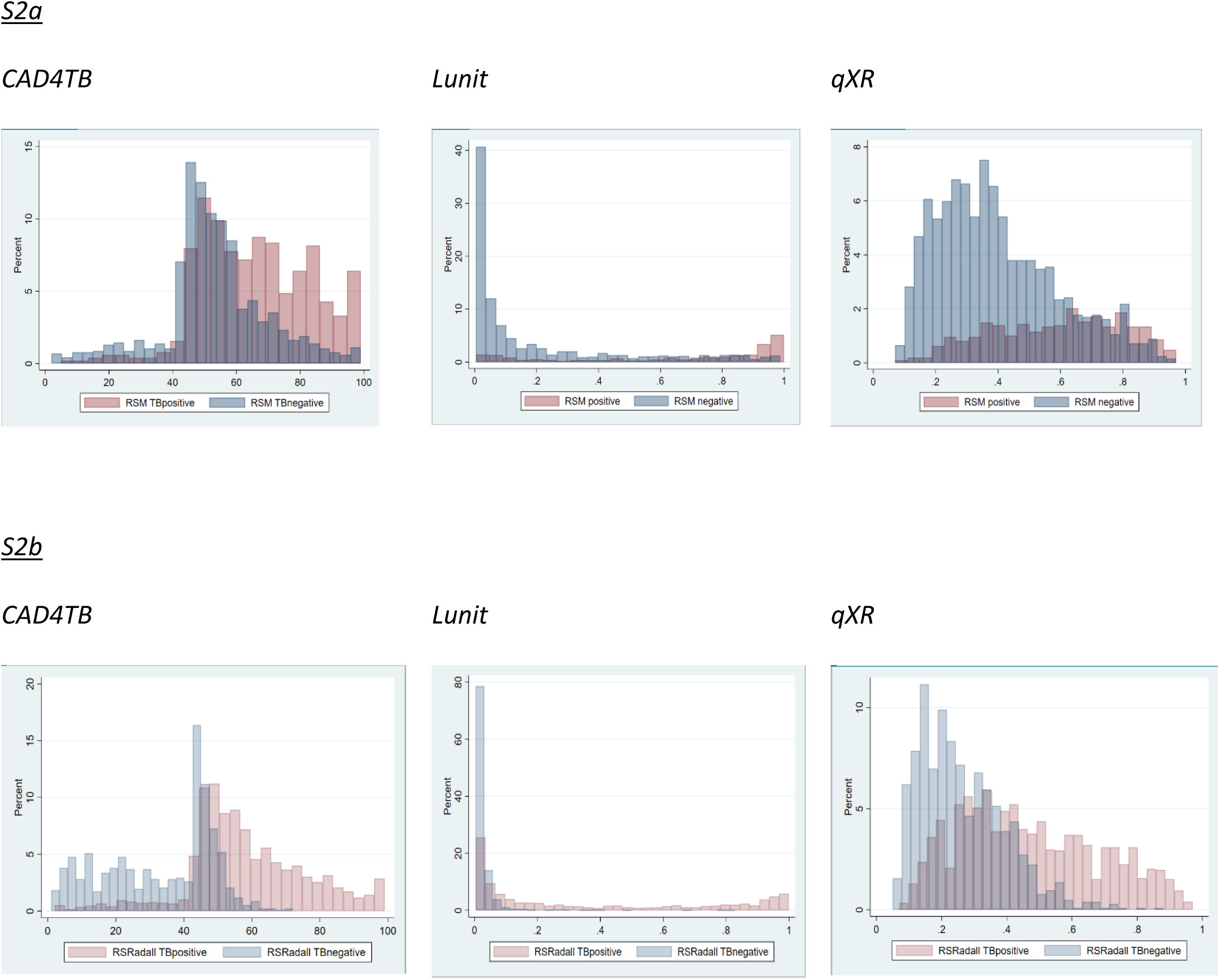
Two-way histogram distribution of abnormality scores from three CAD systems for a) MRS analysis population, and b) RadRS analysis population.

**S3 Figure:**
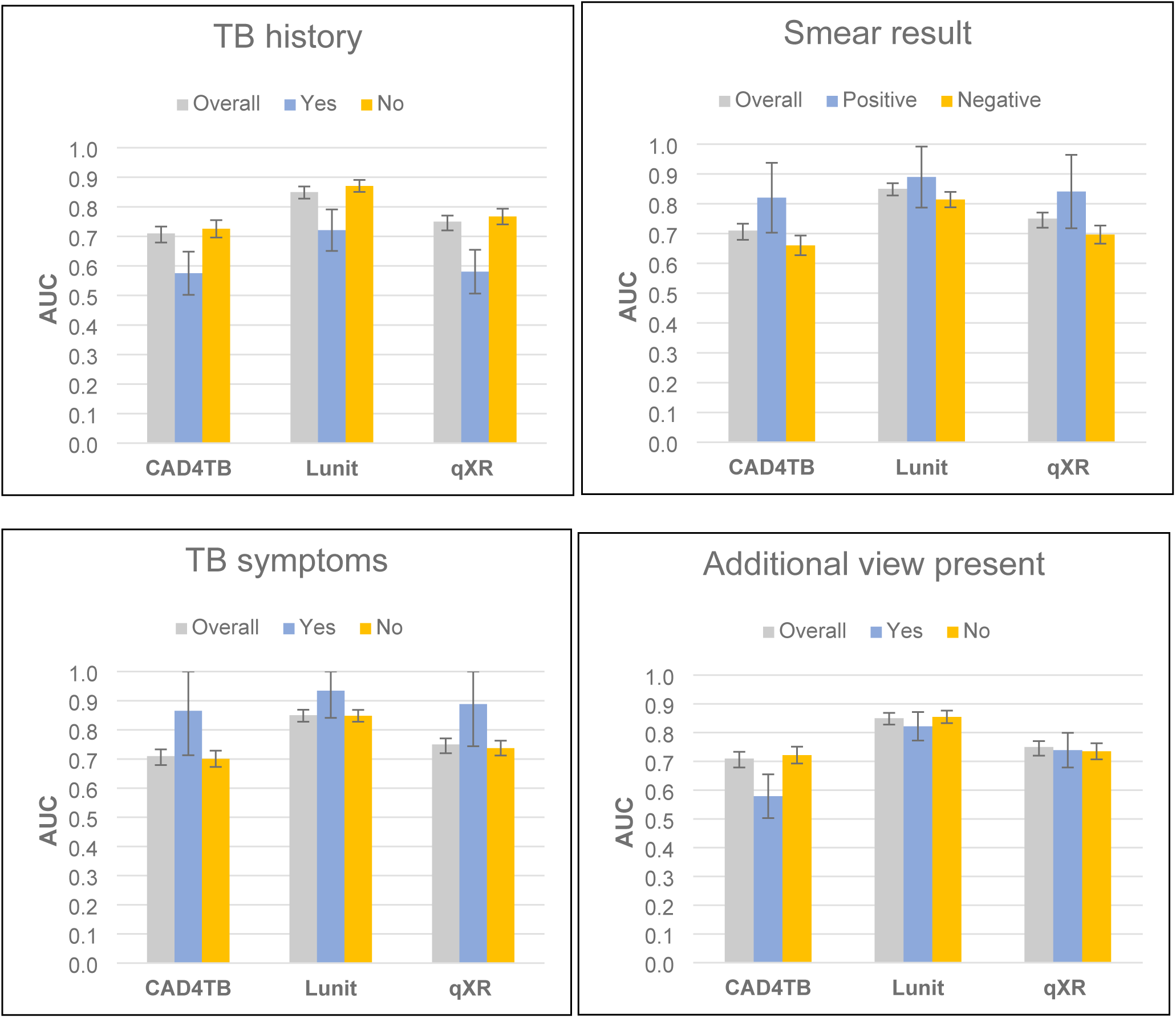
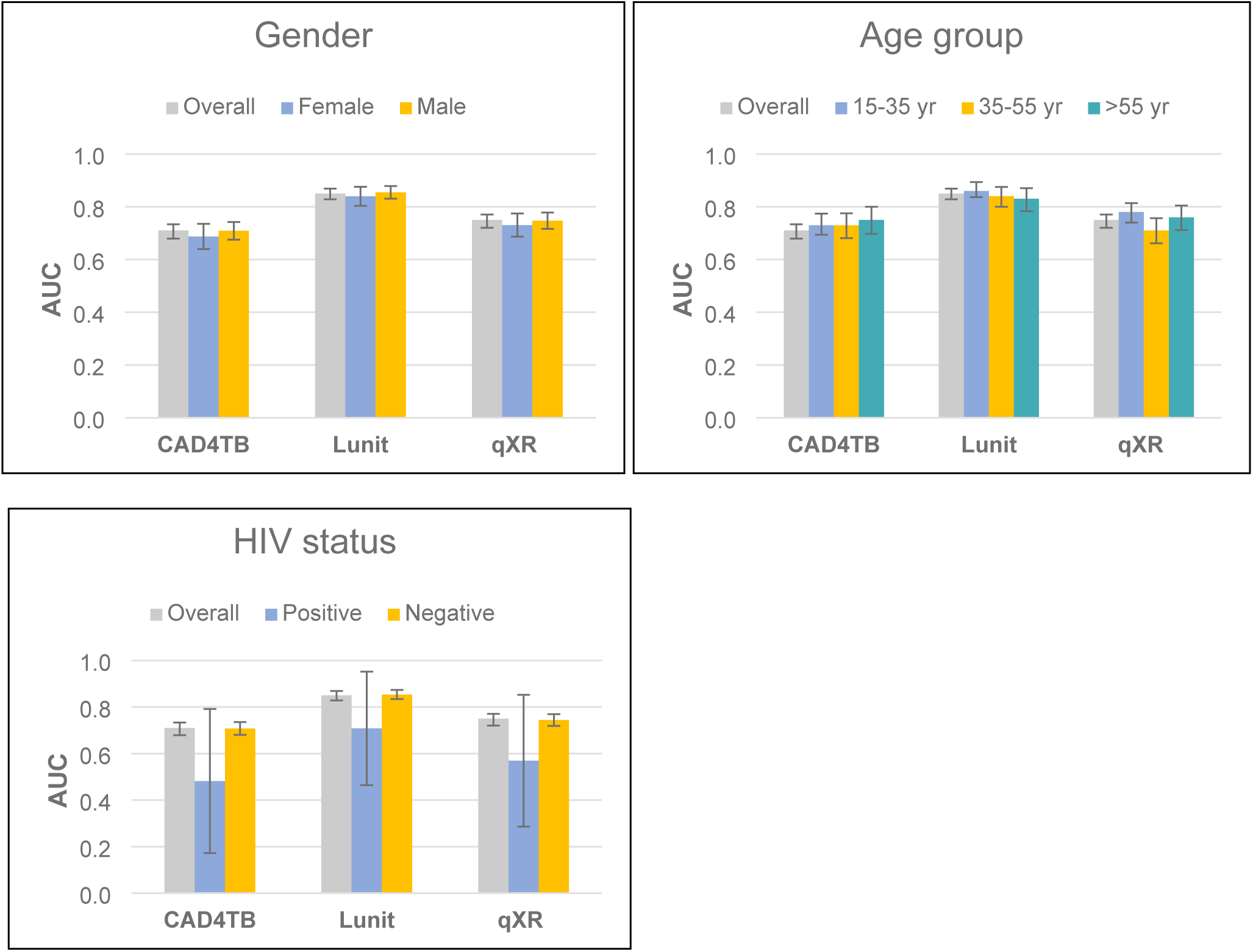
Diagnostic accuracy of three CAD systems across population subgroups.

**S1 Table:** Distribution of study participants by across TB screening countries (MRS analysis population).

| Country (number of Health<br>Assessment Locations) | TB<br>n (%) | non-TB<br>n (%) | Total<br>n (%) |
| --- | --- | --- | --- |
| Bangladesh (2) | 8(1.5%) | 23(1.9%) | 31(1.8%) |
| Burundi | 5(0.9%) | 11(0.9%) | 16(0.9%) |
| Cambodia | 30(5.6%) | 53(4.3%) | 83(4.7%) |
| Chad | 1(0.2%) | 1(0.1%) | 2(0.1%) |
| Ethiopia (3) | 10(1.9%) | 44(3.6%) | 54(3.1%) |
| Iraq | 1(0.2%) | 4(0.3%) | 5(0.3%) |
| Jordan | 1(0.2%) | 11(0.9%) | 12(0.7%) |
| Kenya (3) | 76(14.3%) | 187(15.1%) | 263(14.9%) |
| Malaysia | 81(15.2%) | 164(13.3%) | 245(13.8%) |
| Nepal (2) | 165(31%) | 336(27.2%) | 501(28.3%) |
| Pakistan (2) | 4(0.8%) | 9(0.7%) | 13(0.7%) |
| Russian Federation | 7(1.3%) | 56(4.5%) | 63(3.6%) |
| Rwanda | 5(0.9%) | 9(0.7%) | 14(0.8%) |
| Tanzania | 12(2.3%) | 22(1.8%) | 34(1.9%) |
| Thailand (3) | 32(6.0%) | 66(5.3%) | 98(5.5%) |
| Uganda (4) | 16(3.0%) | 31(2.5%) | 47(2.7%) |
| Ukraine | 7(1.3%) | 68(5.5%) | 75(4.2%) |
| Vietnam (2) | 72(13.5%) | 141(11.4%) | 213(12.0%) |
| **Total** | **533(100.0%)** | **1236(100.0%)** | **1769(100.0%)** |

**S2 Table:** Distribution of study participants across TB screening countries (RadRS analysis population).

| **Country (number of Health Assessment Locations)** | **TB**<br>**n (%)** | **Non-TB**<br>**n (%)** | **Total**<br>**n (%)** |
| --- | --- | --- | --- |
| Bangladesh (2) | 34(1.9%) | 19(1.8%) | 53(1.9%) |
| Burundi | 19(1.1%) | 5(0.5%) | 24(0.9%) |
| Cambodia | 82(4.6%) | 25(2.4%) | 107(3.8%) |
| Chad | 2(0.1%) | 0(0%) | 2(0.1%) |
| Ethiopia (3) | 52(2.9%) | 56(5.4%) | 108(3.8%) |
| Iraq | 5(0.3%) | 9(0.9%) | 14(0.5%) |
| Jordan | 9(0.5%) | 20(1.9%) | 29(1%) |
| Kenya (3) | 268(15%) | 185(17.9%) | 453(16.1%) |
| Malaysia | 256(14.4%) | 111(10.8%) | 367(13.1%) |
| Nepal (2) | 502(28.2%) | 200(19.4%) | 702(25%) |
| Pakistan (2) | 13(0.7%) | 7(0.7%) | 20(0.7%) |
| Russian Federation | 53(3.0%) | 83(8.1%) | 136(4.8%) |
| Rwanda | 14(0.8%) | 14(1.4%) | 28(1.0%) |
| Tanzania | 35(2.0%) | 23(2.2%) | 58(2.1%) |
| Thailand (3) | 93(5.2%) | 75(7.3%) | 168(6.0%) |
| Uganda (4) | 54(3.0%) | 26(2.5%) | 80(2.8%) |
| Ukraine | 81(4.5%) | 104(10.1%) | 185(6.6%) |
| Vietnam (2) | 209(11.7%) | 69(6.7%) | 278(9.9%) |
| **Total** | **1781(100.0%)** | **1031(100.0%)** | **2812(100.0%)** |

**S3 Table:**
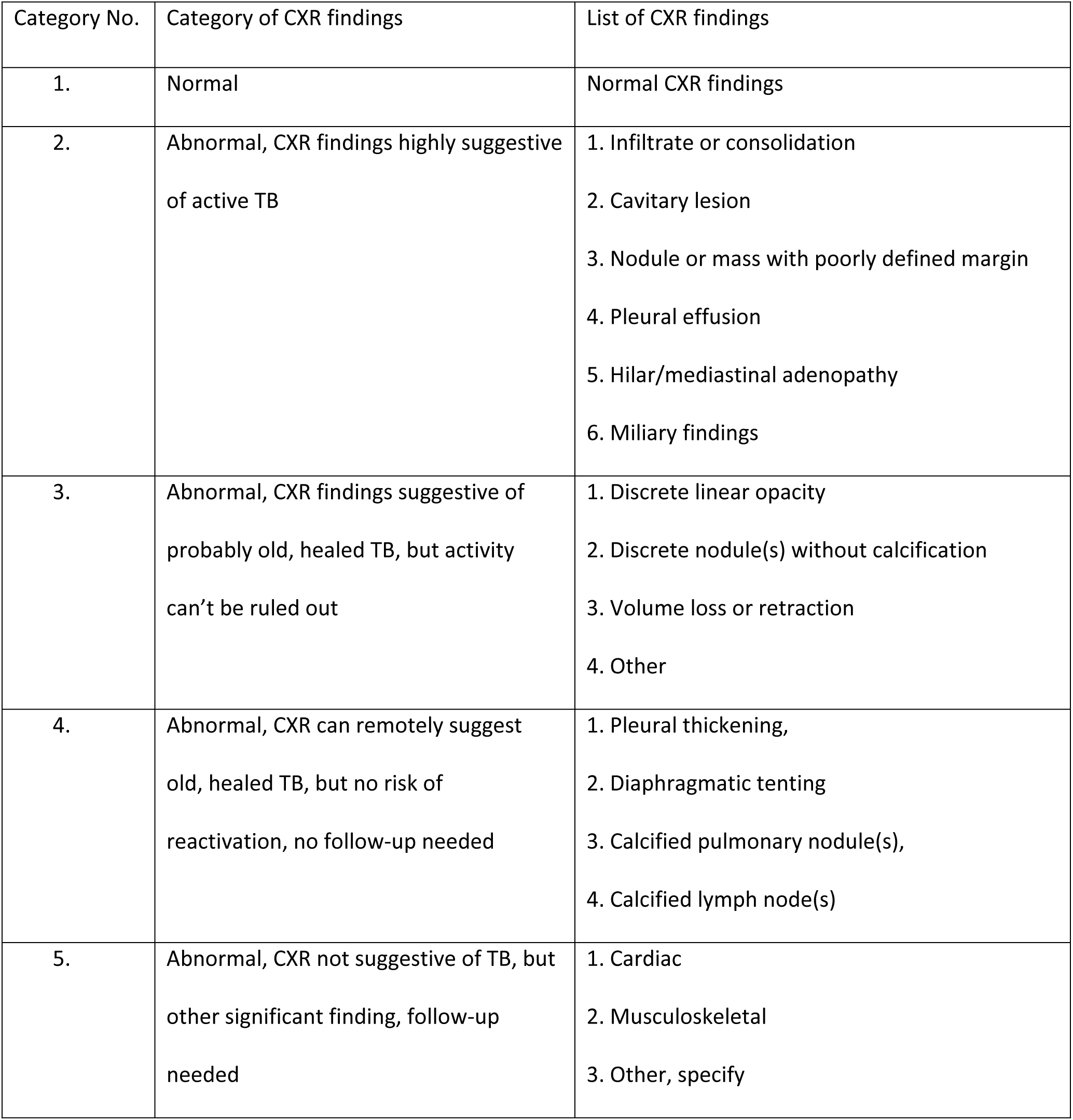
Chest X-ray classifications used by IOM radiologists for detecting pulmonary tuberculosis as part of the DS-3030 CXR reporting form.

| Category No. | Category of CXR findings | List of CXR findings |
| --- | --- | --- |
| 1. | Normal | Normal CXR findings |
| 2. | Abnormal, CXR findings highly suggestive of active TB | 1. Infiltrate or consolidation<br>2. Cavitary lesion<br>3. Nodule or mass with poorly defined margin<br>4. Pleural effusion<br>5. Hilar/mediastinal adenopathy<br>6. Miliary findings |
| 3. | Abnormal, CXR findings suggestive of probably old, healed TB, but activity can't be ruled out | 1. Discrete linear opacity<br>2. Discrete nodule(s) without calcification<br>3. Volume loss or retraction<br>4. Other |
| 4. | Abnormal, CXR can remotely suggest old, healed TB, but no risk of reactivation, no follow-up needed | 1. Pleural thickening,<br>2. Diaphragmatic tenting<br>3. Calcified pulmonary nodule(s),<br>4. Calcified lymph node(s) |
| 5. | Abnormal, CXR not suggestive of TB, but other significant finding, follow-up needed | 1. Cardiac<br>2. Musculoskeletal<br>3. Other, specify |

**S4 Table:** Baseline demographics and clinical characteristics (RadRS analysis population).

| **Variables** | **TB (%)** | **non-TB (%)** | **Total (%)** |
| --- | --- | --- | --- |
| **Total (%)** | 1781 (63.3%) | 1031 (36.7%) | 2812 (100.0%) |
| Male | 1081 (60.7%) | 474 (46.0%) | 1555 (55.3%) |
| Female | 700 (39.3%) | 557 (54.0%) | 1257 (44.7%) |
| **Age group** |  |  |  |
| 15-35 year | 637 (35.8%) | 639 (62.0%) | 1276 (45.4%) |
| 36-55 year | 566 (31.8%) | 277 (26.9%) | 843 (30.0%) |
| 56+ year | 578 (32.5%) | 115 (11.2%) | 693 (24.6%) |
| **Region** |  |  |  |
| Africa | 444 (24.9%) | 309 (30.0%) | 753 (26.8%) |
| Asia Pacific | 1188 (66.7%) | 506 (49.1%) | 1694 (60.2%) |
| Middle East | 14 (0.8%) | 29 (2.8%) | 43 (1.5%) |
| Eastern Europe | 134 (7.5%) | 187 (18.1%) | 321 (11.4%) |
| not indicated | 1 (0.1%) | 0 (0.0%) | 1 (0.0%) |
| **TB symptoms** |  |  |  |
| Cough (any duration) | 23 (1.3%) | 2 (0.2%) | 25 (0.9%) |
| Fever | 7 (0.4%) | 0 (0.0%) | 7 (0.2%) |
| Weight loss | 13 (0.7%) | 3 (0.3%) | 16 (0.6%) |
| Night sweats | 5 (0.3%) | 0 (0.0%) | 5 (0.2%) |
| TB symptoms present $\geq 1$ | 29 (1.6%) | 5 (0.5%) | 34 (1.2%) |
| **Risk groups** |  |  |  |
| History of TB | 260 (14.6%) | 9 (0.9%) | 269 (9.6%) |
| Migrant type (refugee) | 1037 (58.2%) | 577 (56.0%) | 1614 (57.4%) |
| HIV positive | 16 (0.9%) | 3 (0.3%) | 19 (0.7%) |
| **Image quality issue** |  |  |  |
| Yes | 328 (18.4%) | 138 (13.4%) | 466 (16.6%) |
| No | 1453 (81.6%) | 893 (86.6%) | 2346 (83.4%) |
| **Additional view present** |  |  |  |
| Yes | 387 (21.7%) | 124 (12.0%) | 511 (18.2%) |
| No | 1394 (78.3%) | 907 (88.0%) | 2301 (81.8%) |
| **Smear result** |  |  |  |
| Positive | 192 (11.0%) | 6 (2.9%) | 198 (10.2%) |
| Negative | 1546 (89.0%) | 199 (97.1%) | 1745 (89.8%) |

**S5 Table:** Chest X-ray and microbiological test results (RadRS analysis population).

| **Variables** | ***TB (%)*** | ***non-TB (%)*** | ***Total RadRS (%)*** |
| --- | --- | --- | --- |
| **Total (%)** | *1781 (63.3%)* | *1031 (36.7%)* | *2812 (100.0%)* |
| **CXR findings Interpretation results** |  |  |  |
| 1=Normal | 0 (0.0%) | 893 (86.6%) | 893 (31.8%) |
| 2=Abnormal CXR, highly suggestive of active TB (follow-up required) | 1310 (73.6%) | 0 (0.0%) | 1310 (46.6%) |
| 3=Abnormal CXR, may suggest old, healed TB but active TB can't be ruled out (follow-up required) | 440 (24.7%) | 0 (0.0%) | 440 (15.6%) |
| 4=Abnormal CXR, can remotely suggest old, healed TB but minimal risk of reactivation (NO follow-up required) | 0 (0.0%) | 134 (13.0%) | 134 (4.8%) |
| 5=Abnormal CXR, not suggestive of TB (follow-up required) | 31 (1.7%) | 4 (0.4%) | 35 (1.2%) |
| 6=Other abnormalities (No follow-up required) | 0 (0.0%) | 0 (0.0%) | 0 (0.0%) |
| **Bacteriology result [for those done =<14 days CXR to culture date]** |  |  |  |
| Culture +, Xpert MTB/RIF not done | 486 (27.3%) | 11 (1.1%) | 497 (17.7%) |
| Culture +, Xpert MTB/RIF + | 88 (4.9%) | 2 (0.2%) | 90 (3.2%) |
| Culture +, Xpert MTB/RIF - | 2 (0.1%) | 0 (0.0%) | 2 (0.1%) |
| Culture -, Xpert MTB/RIF + | 1 (0.1%) | 0 (0.0%) | 1 (0.0%) |
| Culture -, Xpert MTB/RIF not done | 1155 (64.9%) | 190 (18.4%) | 1345 (47.8%) |
| Culture -, Xpert MTB/RIF - | 9 (0.5%) | 2 (0.2%) | 11 (0.4%) |
| Culture not done, Xpert MTB/RIF not done | 40 (2.3%) | 826 (80.1%) | 866 (30.8%) |

**S6 Table:** Estimated diagnostic accuracy of CAD systems (RadRS analysis population)

| **Sensitivity and specificity category** | **CAD System** | **Sensitivity (95% CI)** | **Specificity (95% CI)** | **Threshold score** |
| --- | --- | --- | --- | --- |
| Specificity at 90% sensitivity and associated threshold score | CAD4TB | 91.4 (90–92.7) | 54.6 (51.3–57.8) | 43 |
|  | Lunit | 90.0 (88.5–91.4) | 45.5 (42.4–48.6) | 0.014 |
|  | qXR | 91.0 (89.5–92.3) | 37.5 (34.6–40.6) | 0.2 |
| Sensitivity at 70% specificity and associated threshold score | CAD4TB | 84.1 (82.3–85.8) | 70.9 (67.9–73.8) | 46 |
|  | Lunit | 80.9 (78.9–82.7) | 70.0 (67.1–72.8) | 0.024 |
|  | qXR | 70.0 (67.8–72.1) | 70.6 (67.7–73.4) | 0.33 |
| Sensitivity and specificity at manufacturer recommended threshold score | Lunit, low threshold, high sensitivity | 53.1 (50.8–55.5) | 98.4 (97.5–99.1) | 0.150 |
|  | Lunit, middle threshold | 42.9 (40.6–45.2) | 99.2 (98.5–99.7) | 0.300 |
|  | Lunit, high threshold, high specificity | 36.6 (34.3–38.8) | 98.4 (97.5–99.1) | 0.450 |
|  | qXR, routine TB threshold | 35.0 (32.8–37.2) | 96.6 (95.3–97.6) | 0.55 |
|  | qXR, high risk TB threshold | 13.8 (12.2–15.4) | 99.7 (99.2–99.9) | 0.75 |

*Lunit score was 6 decimal places, but it was rounded to three decimals.*

**S7 Table:**
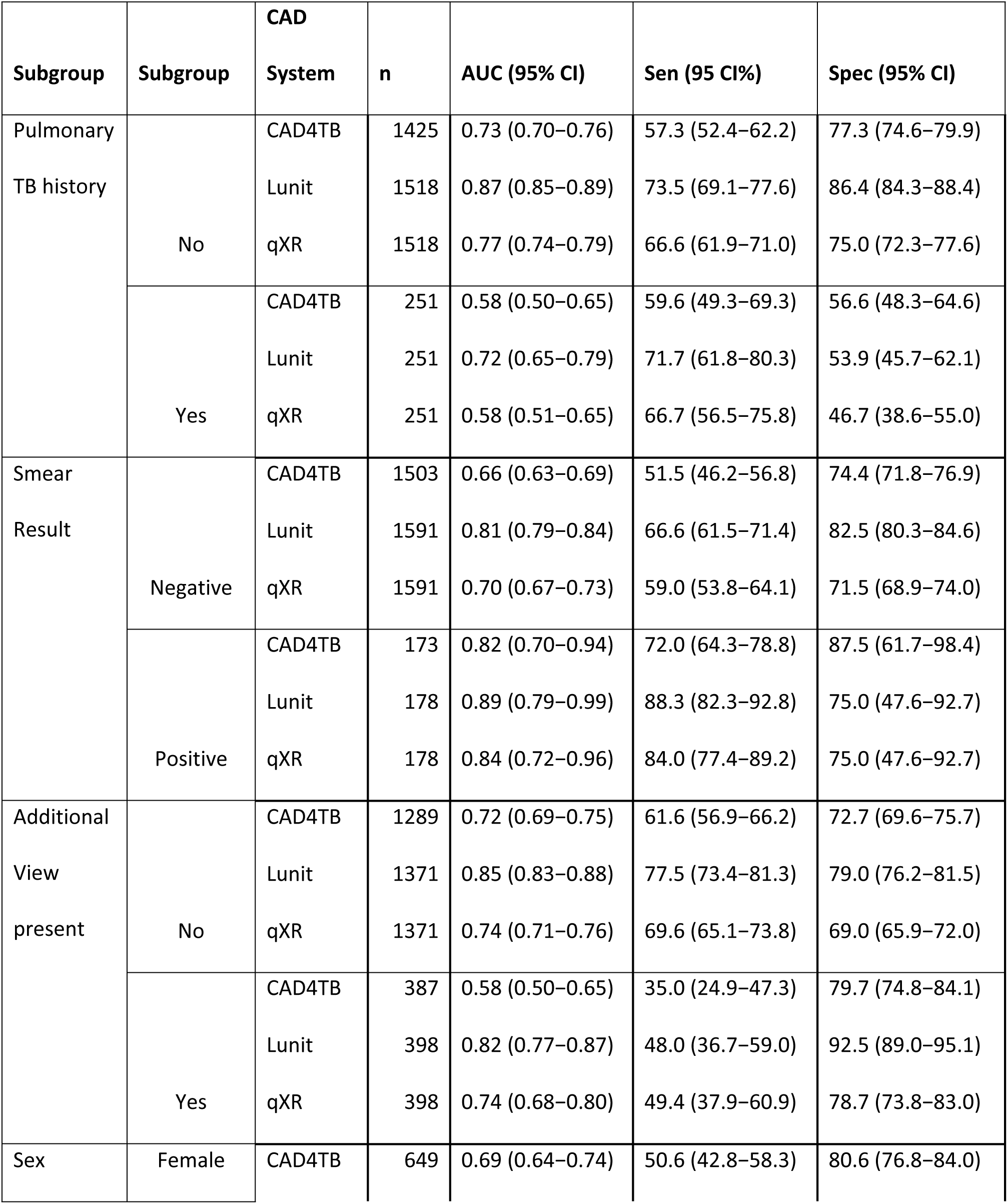
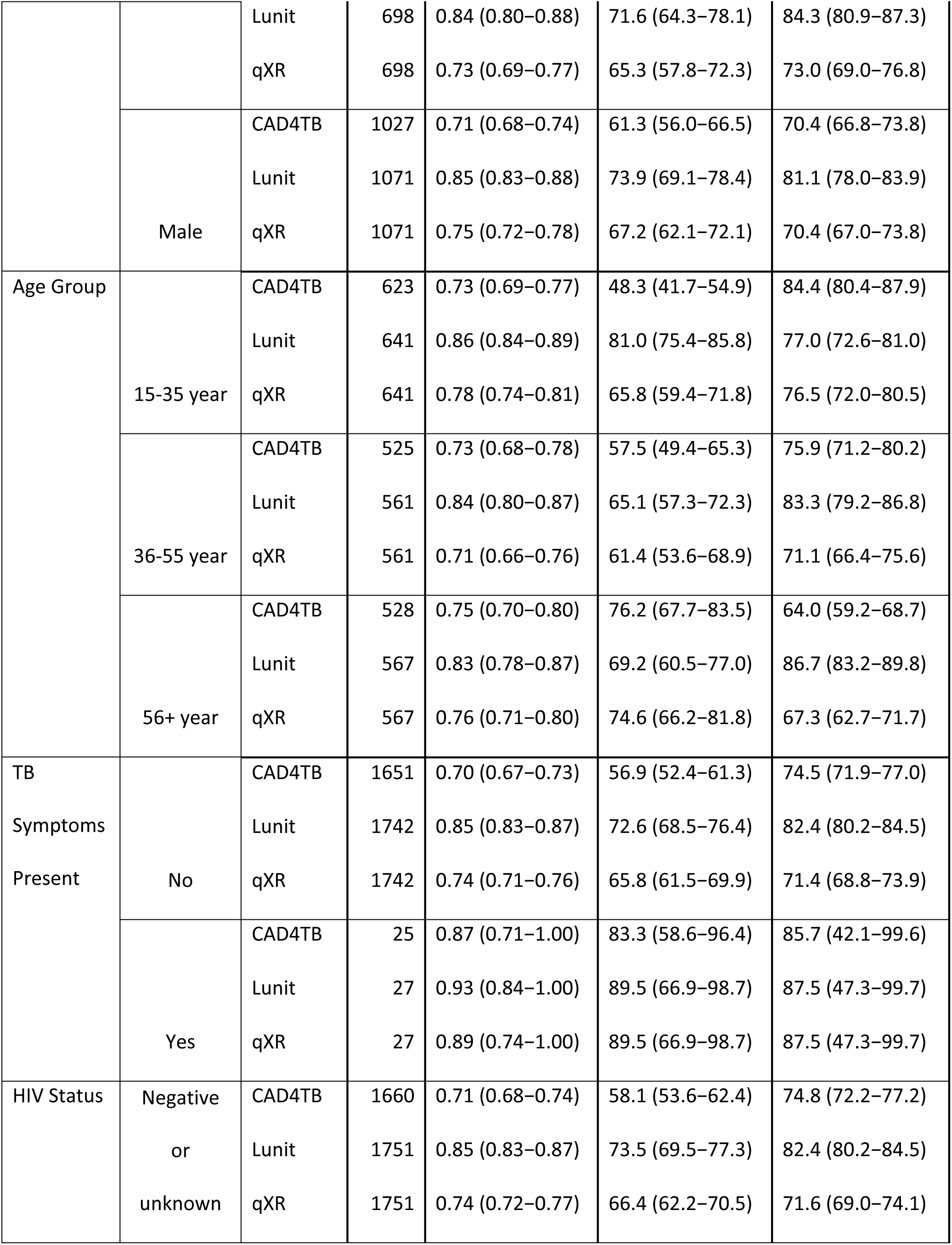
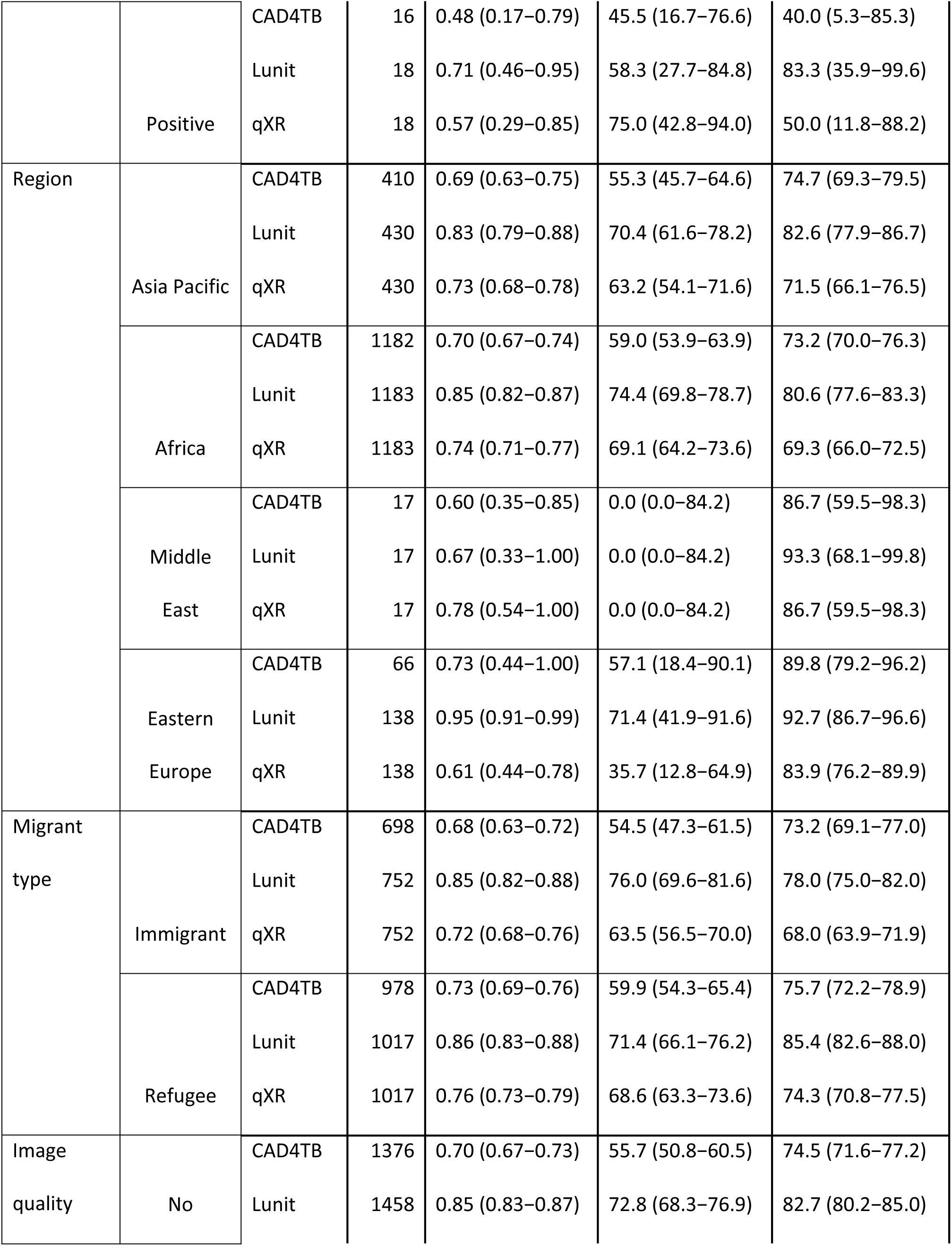
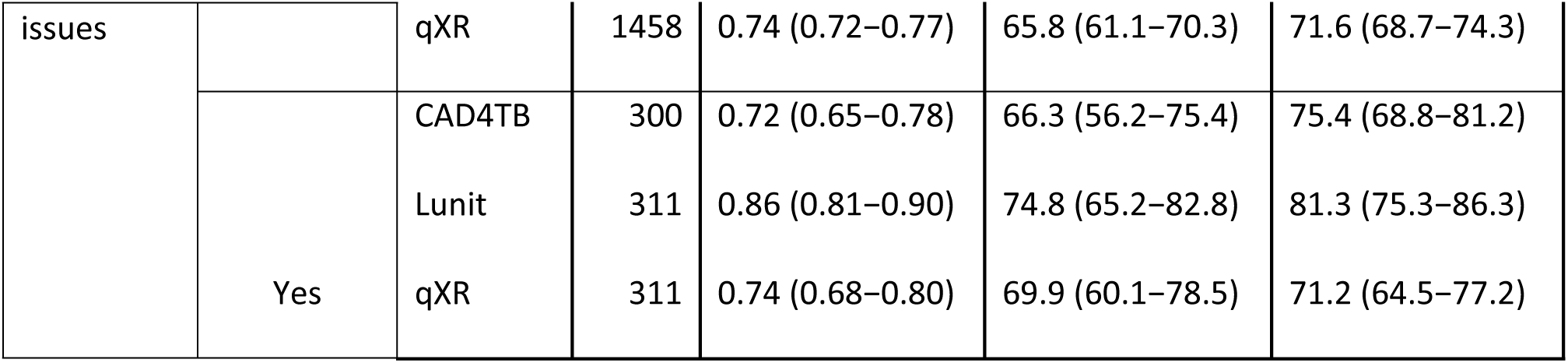
Accuracy of the CAD systems in different population subgroups (MRS analysis population)

